# Time-resolved dynamic optical coherence tomography for retinal blood flow analysis

**DOI:** 10.1101/2023.10.31.23297800

**Authors:** Philippe Valmaggia, Philippe C. Cattin, Robin Sandkühler, Nadja Inglin, Tilman P. Otto, Silke Aumann, Michel M. Teussink, Richard F. Spaide, Hendrik P. N. Scholl, Peter M. Maloca

**Affiliations:** Department of Biomedical Engineering, University of Basel, 4123 Allschwil, Switzerland; Institute of Molecular and Clinical Ophthalmology Basel (IOB), Basel, 4031 Basel, Switzerland; Department of Ophthalmology, University Hospital Basel, 4031 Basel, Switzerland; Heidelberg Engineering GmbH, 69115 Heidelberg, Germany; Vitreous Retina Macula Consultants of New York, NY 10022, USA

**Author notes:** Corresponding author: Philippe Valmaggia, Department of Biomedical Engineering, University of Basel, 4123 Allschwil, Switzerland.

**Keywords:** time-resolved, dynamic, optical coherence tomography, retinal blood flow, flow velocity

## Abstract

**Purpose:** Optical coherence tomography (OCT) representations in clinical practice are static and do not allow for a dynamic visualisation and quantification of blood flow. This study aims to present a method to analyse retinal blood flow dynamics using time-resolved structural optical coherence tomography (OCT).

**Methods:** We developed novel imaging protocols to acquire video-rate time-resolved OCT B-scans (1024 x 496 pixels, 10° field of view) at four different sensor integration times (integration time of 44.8 μs at a nominal A-scan rate of 20 kHz, 22.4 μs at 40 kHz, 11.2 μs at 85 kHz, 7.24 μs at 125 kHz). The vessel centres were manually annotated for each B-scan and surrounding subvolumes were extracted. We used a velocity model based on signal-to-noise ratio (SNR) drops due to fringe washout to calculate blood flow velocity profiles in vessels within five optic disc diameters of the optic disc rim.

**Results:** Time-resolved dynamic structural OCT revealed pulsatile SNR changes in the analysed vessels and allowed the calculation of potential blood flow velocities at all integration times. Fringe washout was stronger in acquisitions with longer integration times; however, the ratio of the average SNR to the peak SNR inside the vessel was similar across all integration times.

**Conclusions:** We demonstrated the feasibility of estimating blood flow profiles based on fringe washout analysis, showing pulsatile dynamics in vessels close to the optic nerve head using structural OCT. Time-resolved dynamic OCT has the potential to uncover valuable blood flow information in clinical settings.

**Commercial relationships:** PV received funding from the Swiss National Science Foundation (Grant 323530_199395), the Janggen-Pöhn Stiftung and AlumniMedizin Basel and discloses personal compensation from Heidelberg Engineering GmbH. TPO, SA and MMT are salaried employees of Heidelberg Engineering GmbH, 69115 Heidelberg, Germany. RFS discloses personal compensation from Topcon Medical Systems, Roche, Bayer, Heidelberg Engineering and Genentech. HPNS is supported by the Swiss National Science Foundation (Project funding: “Developing novel outcomes for clinical trials in Stargardt disease using structure/function relationship and deep learning” #310030_201165, and National Center of Competence in Research Molecular Systems Engineering: “NCCR MSE: Molecular Systems Engineering (phase II)” #51NF40-182895), the Wellcome Trust (PINNACLE study), and the Foundation Fighting Blindness Clinical Research Institute (ProgStar study). HPNS is a member of the Scientific Advisory Board of Boehringer Ingelheim Pharma GmbH & Co; Claris Biotherapeutics Inc.; Eluminex Biosciences; Gyroscope Therapeutics Ltd.; Janssen Research & Development, LLC (Johnson & Johnson); Novartis Pharma AG (CORE); Okuvision GmbH; ReVision Therapeutics Inc.; and Saliogen Therapeutics Inc. HPNS is a consultant of: Alnylam Pharmaceuticals Inc.; Gerson Lehrman Group Inc.; Guidepoint Global, LLC; and Intergalactic Therapeutics Inc. HPNS is member of the Data Monitoring and Safety Board/Committee of Belite Bio (CT2019-CTN-04690-1), F. Hoffmann-La Roche Ltd (VELODROME trial, NCT04657289; DIAGRID trial, NCT05126966; HUTONG trial) and member of the Steering Committee of Novo Nordisk (FOCUS trial; NCT03811561). All arrangements have been reviewed and approved by the University of Basel (Universitätsspital Basel, USB) and the Board of Directors of the Institute of Molecular and Clinical Ophthalmology Basel (IOB), in accordance with their conflict-of-interest policies. Compensation is being negotiated and administered as grants by USB, which receives them on its proper accounts. HPNS is co-director of the Institute of Molecular and Clinical Ophthalmology Basel (IOB), which is constituted as a non-profit foundation and receives funding from the University of Basel, the University Hospital Basel, Novartis and the government of Basel-Stadt. PMM is a consultant of Roche and holds intellectual properties for machine learning at MIMO AG and VisionAI, Switzerland. Funding organisations had no influence on the design, performance or evaluation of the current study. The other authors declare no conflict.

## Introduction

Optical coherence tomography (OCT) is a technology that enables the visualisation of retinal structures on a micrometre scale.^1,2^ Its benefits of being non-invasive, fast and easy to operate have made OCT become one of the most widely used imaging techniques, particularly in ophthalmology.^3^ Since its first demonstration in 1991, OCT has been enhanced and nowadays, allows the visualisation of intravascular flow regions through OCT angiography (OCTA).^4^ OCTA is based on the detection of OCT signal changes over time and allows the visualisation of chorioretinal vasculature down to the microvasculature in an unprecedented way.

However, the clinical representations of both OCT and OCTA are static. For visualising flow dynamics and analysing changes within single vessels over time, ophthalmologists and researchers must rely on other acquisition modalities such as Doppler OCT, laser speckle flowgraphy, the retinal function imager or variable interscan time analysis (VISTA) in macular OCTA.^5–13^ These techniques are mostly used in specialised research centres and are not yet widely used in everyday clinical use. Due to the relatively high demands on technology and experience, they have not yet been implemented in a routine clinical application that can be used on a daily basis. However, the scan patterns in OCT devices are generally customisable and modifications would allow for dynamic acquisitions and visualisations in clinical settings. Sequentially acquired OCT signals at the same position particularly fluctuate within and in close proximity to vessels; accordingly, they have the greatest contribution to OCTA signal generation.^4,14^

The OCT signals are generated by measuring the interference pattern of reflected light waves from a sample and a reference mirror.^1,15,16^ The reflected light waves interfere constructively or destructively, generating interference patterns of light and dark bands which are called interference fringes^15,17^ – appearing similar to the edge of a fabric fringe. When the sample or instrument moves during image acquisition, the phase of the reflected light changes. This modulation of the interference can cause the interference fringes to vanish, a phenomenon called fringe washout. The fringe washout leads to a signal-to-noise ratio (SNR) drop in the image, which tends to be larger in case of a larger sample motion.^18–20^ This can occur at higher sample velocities or increased sensor integration times.^18^ Taking advantage of the knowledge that fringe washout is directly linked to the velocity of the sample motion, we investigated the possibility of calculating flow profiles in time-resolved structural OCT data. An overview of the general principle of fringe washout can be found in Figure 1.

**Figure 1.**
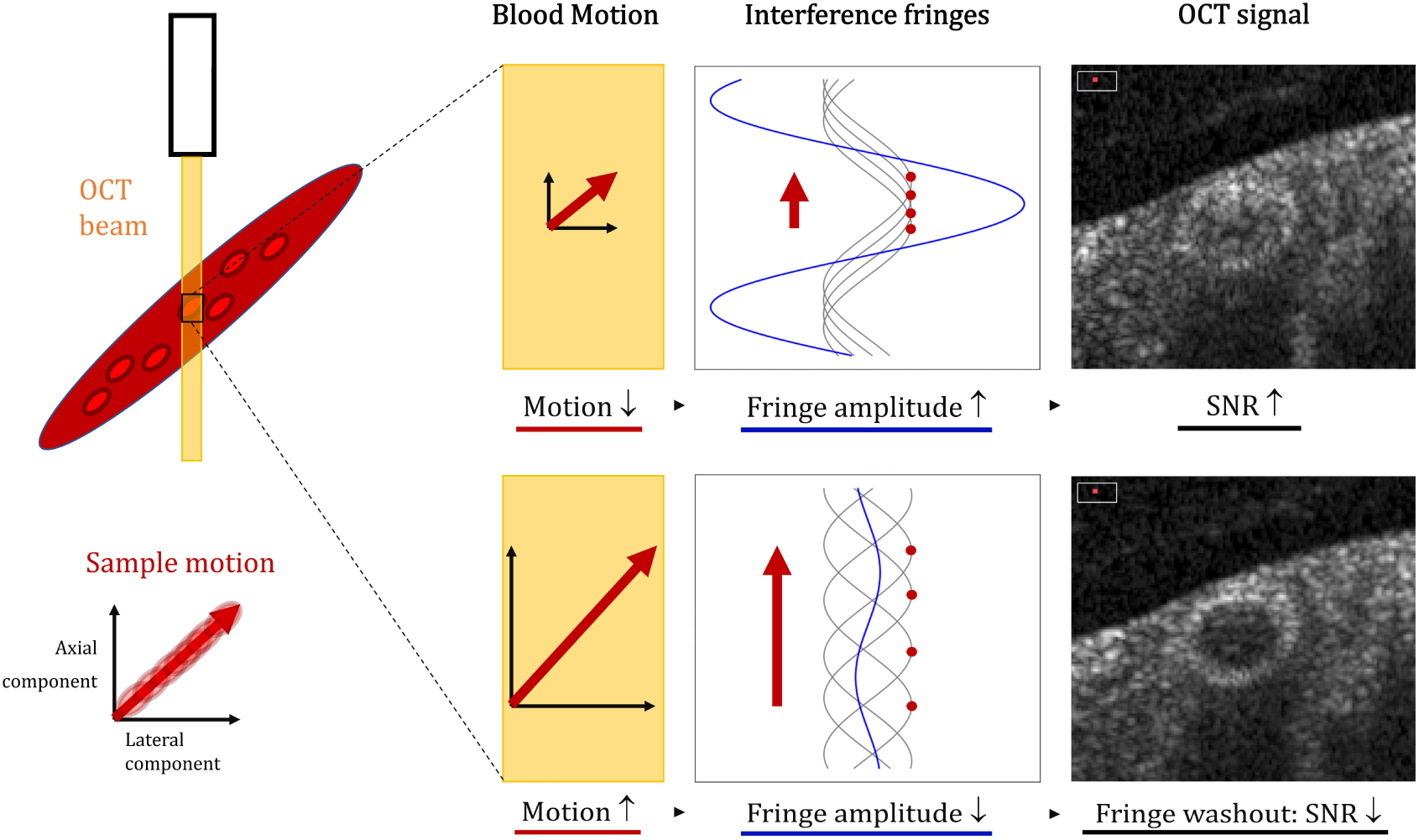
Illustration of the fringe washout in spectral-domain optical coherence tomography (OCT). Fringe washout describes the signal drop in the OCT when the sample moves through the sample beam during illumination. The sample motion can be split into two components: a lateral component and an axial component. When the integration time is shorter or the velocity is smaller, there is less motion, the fringe amplitude is higher and, hence, the OCT intensity is higher. When the integration time is longer or the velocity is higher, the sample has a larger motion, which leads to a lower fringe amplitude. This fringe washout causes a higher SNR drop and a lower OCT intensity (bottom right).

To investigate blood flow velocity, we modified the OCT scan patterns of a commercially available device to allow for time-resolved dynamic acquisitions. Furthermore, we analysed these images regarding the fringe washout and present dynamically calculated blood flow profiles.

## Methods

### Data acquisition

Data were acquired at the Augenarztpraxis Dr. Maloca in Lucerne, Switzerland. The study was performed according to the Declaration of Helsinki, and ethics approval was granted by the Ethics Committee of Northwestern and Central Switzerland (EKNZ 2021-02360). Written informed consent was obtained from all participants.

The images were acquired with a Heidelberg Spectralis OCT with an investigational acquisition module. The OCT volume acquisition pattern was modified to continuous B-scans with a 10° field of view without OCT tracking through the on-board dual-beam confocal scanning laser ophthalmoscope tracking system. This prevented irregular OCT B-scan acquisition due to compensatory OCT-beam steering by the tracking system, which has a variable time lag. The patients were informed about the acquisition procedure to ensure good image quality. The subjects were instructed to fixate on the foveal fixation target, close their eyes, blink once and then leave their eyes open. Acquisitions were performed for several seconds, where a manual press on the joystick of the OCT device allowed to start and end the acquisition. In the case of blinks or major ocular movements, the process was repeated.

The acquisitions were performed between the optic nerve head centre and five optic disc diameters (ODDs) from the optic nerve rim and graded by eccentricity 0–6 (0: optic nerve head centre, 1: neuroretinal rim, 2: 1 ODD superotemporal to the neuroretinal rim, 3: 2 ODDs, 4: 3 ODDs, 5: 4 ODDs, 6: 5 ODDs). The major visible vessels were imaged perpendicularly, so that the cross-section was visible.

All time-resolved acquisitions were performed with continuous B-scans, each single scan comprising 1024 A-scans with 496 pixels, with a 10° field of view. This allowed the same B-scan to be repeated at the same location with a high sampling density over a longer period of time and still meet the requirements regarding light safety. All locations were acquired at four different A-scan integration times 44.8, 22.4, 11.2 and 7.24 μs, corresponding to nominal A-scan rates of 20, 40, 85 and 125 kHz, respectively. This enabled video-rate sequential B-scan acquisitions. Each separate B-scan was saved as a single image without averaging. Notably, the SNR in spectral-domain (SD) OCTs depends on the integration time according to the following established formula^21–23^:

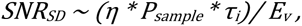

where *η* corresponds to the spectrometer efficiency, *P_sample_* to the power returned in the sample arm, *τ_i_* to the integration time and *E_v_* to the energy per photon. A validation of this formula was investigated during this in vivo study. The SNR for each individual B-scan was calculated with the raw data by the ratio of the maximum intensity to the noise level. Overall, the SNR values were compared for each integration time.

### Image processing

All image stacks were exported from the OCT device in normalised raw format (.vol files, image intensities normalised between 0 and 1 at a B-scan level). In the first step, all image stacks were transformed for a visual representation by multiplying all image intensities by 255 times the 4^th^ square root of the normalised intensity. These transformed images stacks were then registered using a rigid body transformation model with a pyramid processing scheme.^24^ This registration pipeline was adapted to reduce the effect of speckle noise in the registration process. The mean of the first 10 images of the image stack was used as the reference image for the registration of the complete image stack. Rigid body registration was applied to prevent distortion of the retina and to allow visualisation of the time-resolved B-scan stack. In the second step, the vessel centres of prominently visible vessels were manually annotated, and subvolumes of 7 × 7 pixels around the centre were extracted from each B-scan. This subvolume corresponded to physical dimensions of 20 µm along the B-scan axis and 27 µm along the A-scan axis.

In a second step, the extracted subvolumes were analysed with their original, non-normalised intensities, which were obtained from the metadata of the .vol files. The SNR in the individual vessel subvolumes were then calculated as the ratio of the average pixel intensity in the subvolume to the noise level of the B-scan, assuming shot noise limited detection. The noise level of the B-scan was approximated using the Heidelberg Quality score (in dB) and the maximum intensity value of the B-Scan. A depth-specific noise level to account for roll-off was not obtainable with the present data. An overview of the main image processing steps is shown in Figure 2.

**Figure 2.**
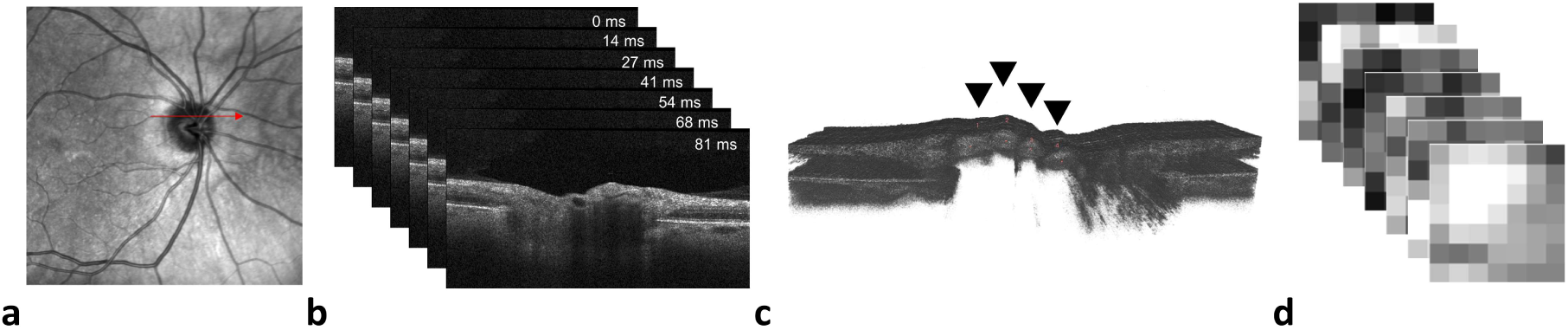
Time-resolved dynamic optical coherence tomography-based extraction of vessel subvolumes. **a** Scanning laser ophthalmoscopy (SLO) image. The red arrow indicates the position of the continuous acquisitions, which can be started by a press on the joystick. **b** The time-resolved B-scans are continuously acquired with a time stamp for each B-scan. **c** The continuous images are registered and reconstructed as a three-dimensional volume, where the third dimension represents the time axis. On each B-scan, the vessel centres were manually annotated. **d** Subvolumes of 7 × 7 pixels surrounding the vessel centre were extracted and further processed for fringe washout analysis.

### Flow profile generation

Fringe washout is a phenomenon occurring when imaged particles are moving which leads to a SNR drop in the OCT image. The effect is linked to the integration time and is attributed to the modulation of the interference signal when particles are in motion. Typically, the fringe washout is more pronounced as the integration time increases. Yun et al. presented formulas for the fringe washout describing separately the axial and lateral components of particle movements.^18^ In the case of oblique motion, the axial component is orders of magnitude larger when compared with the lateral component.^19^ In this work, we used the squared sinc function presented for the axial component of the fringe washout to calculate blood flow velocity profiles^18^:

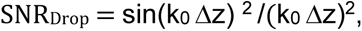

where k_0_ = 2* π / λ and Δz = n * v * τ. k_0_ corresponds to the central wavenumber of the light source, λ to the central wavelength of the light source, Δz to the displacement in the z-axis, n to the refractive index of the eye, τ to the integration time, and v to the velocity.

This quadratic sinc function, with a known SNR drop, was solved numerically for velocity. All numerical solutions per B-scan were stored as a potential flow velocity. The results of the equation for each B-scan, SNR_Drop_, were used to generate the dynamic flow profiles. Figure 3 presents the SNR_Drop_ profiles at the four acquisition rates with calculated velocities in mm/s for any moving object. For this study, the average SNR of the 7 × 7 B-scan subvolume was divided by the maximum SNR at the pixel level of the complete vessel subvolume (7 × 7 × number of B-scans) to calculate the SNR_Drop_. All calculations were made with absolute SNR values. The calculated velocities are presented in arbitrary units since the formula only takes into account the axial component and the SNR inside the vessels without flow, which would be the reference for SNR_Drop_ calculations, could not be determined.

**Figure 3.**
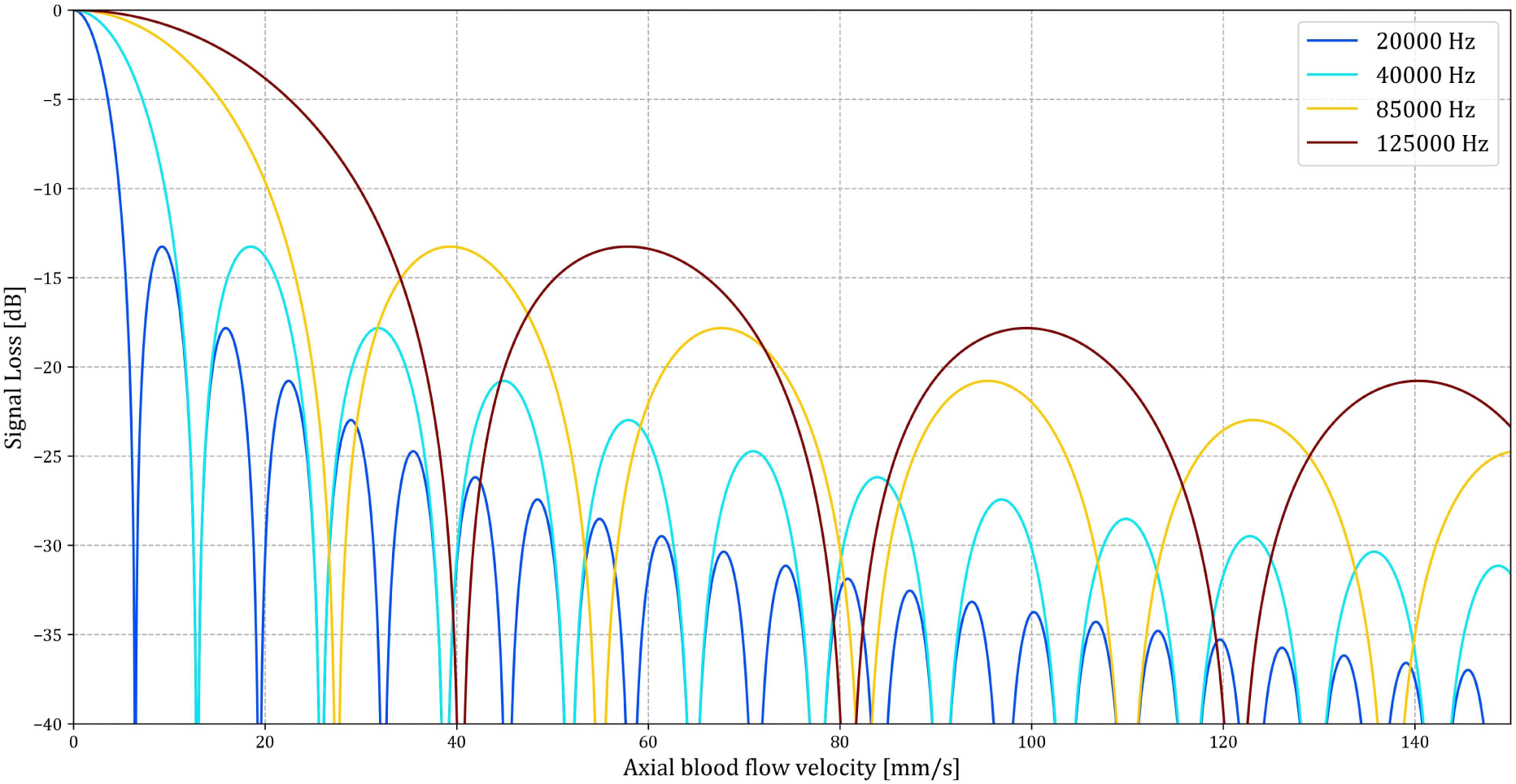
Axial blood flow velocity component calculation. The axial blood flow velocity is calculated at SNR drops for the four nominal acquisition rates of 20, 40, 85 and 125 kHz. As seen, several potential axial blood flow velocities are based on a single signal-to-noise ratio (SNR) drop. The numerical solutions to the equation were used to generate the blood flow profiles in the time-resolved dynamic OCT acquisitions. The values here represent the calculated velocities in mm/s, where the SNR drop is compared against a known reference SNR.

### Data analysis

The generated flow profiles were quantitatively and qualitatively analysed. On a qualitative level, the generated dynamic OCTs and the course of the pulse waves were analysed. The quantitative analysis was focused on the flow profiles, where the minimal, maximal, and average velocities for each vessel at each B-scan were calculated. The minimal velocity refers to the smallest numerical solution of the SNR_Drop_ equation, the maximum velocity refers to the largest solution, and the average velocity refers to the average of all numerical solutions per B-scan.

Furthermore, the exam duration, the SNR of the B-scans, the mean and maximal intensity of the subvolumes, and the maximum image intensity in the raw data were analysed. Programming was performed with Linux shell scripts, Python 3.9 (Python Software Foundation, Wilmington, USA) and R V4.2.2 (Foundation for Statistical Computing, Vienna, Austria). The data were visualised with Python 3.9, R V4.2.2 and 3DSlicer V4.11.^25,26^

## Results

A total of 164 time-resolved image stacks from six healthy subjects were acquired and manually annotated. All acquisitions were performed in the right eyes of the patients. The image stacks consisted of 47 to 483 B-scans, and between 1 and 5 vessels were annotated per volume. In summary, 466 vessels were annotated in the time-series data, totalling 94,619 annotated vessel centres.

### Qualitative analysis

Time-resolved dynamic acquisitions of structural OCT are feasible in healthy subjects and show changes in reconstructed images over time. These changes are of different nature, where pulsatile profiles can be clearly identified. Supplementary Video 1 shows the pulsation inside an artery at the centre of the optic nerve as changes in image intensity. Furthermore, we can see pulsation of the optic nerve head tissue. Supplementary Video 2 shows a longitudinal cut through the same artery, where we can observe the fringe washout along the vessel. We detected an increase in the fringe washout at the centre of the vessel, showing a higher relative flow velocity. In addition, we identified a bimodal peak of the fringe washout.

Qualitative analysis of the generated flow profiles confirmed this pulsatility for the SNR changes and the calculated flow velocities. As depicted for an acquisition at the optic nerve head centre in Figure 4, the flow profiles show pulsatility where maximal and minimal velocities were clearly distinguishable. In addition, the flow velocities showed a rapid increase, followed by a gradual decrease towards a steady-state until the pulsatile cycle restarted. As shown in Figure 5, this flow profile could be visualised at all four acquisition rates, with a constant repetition of approximately four pulse visualisations within 3 s. These observations suggest that the pulsation propagates in correspondence with the cardiac cycle, which corresponds in this example to a heart rate of 80 beats per minute.

**Figure 4.**
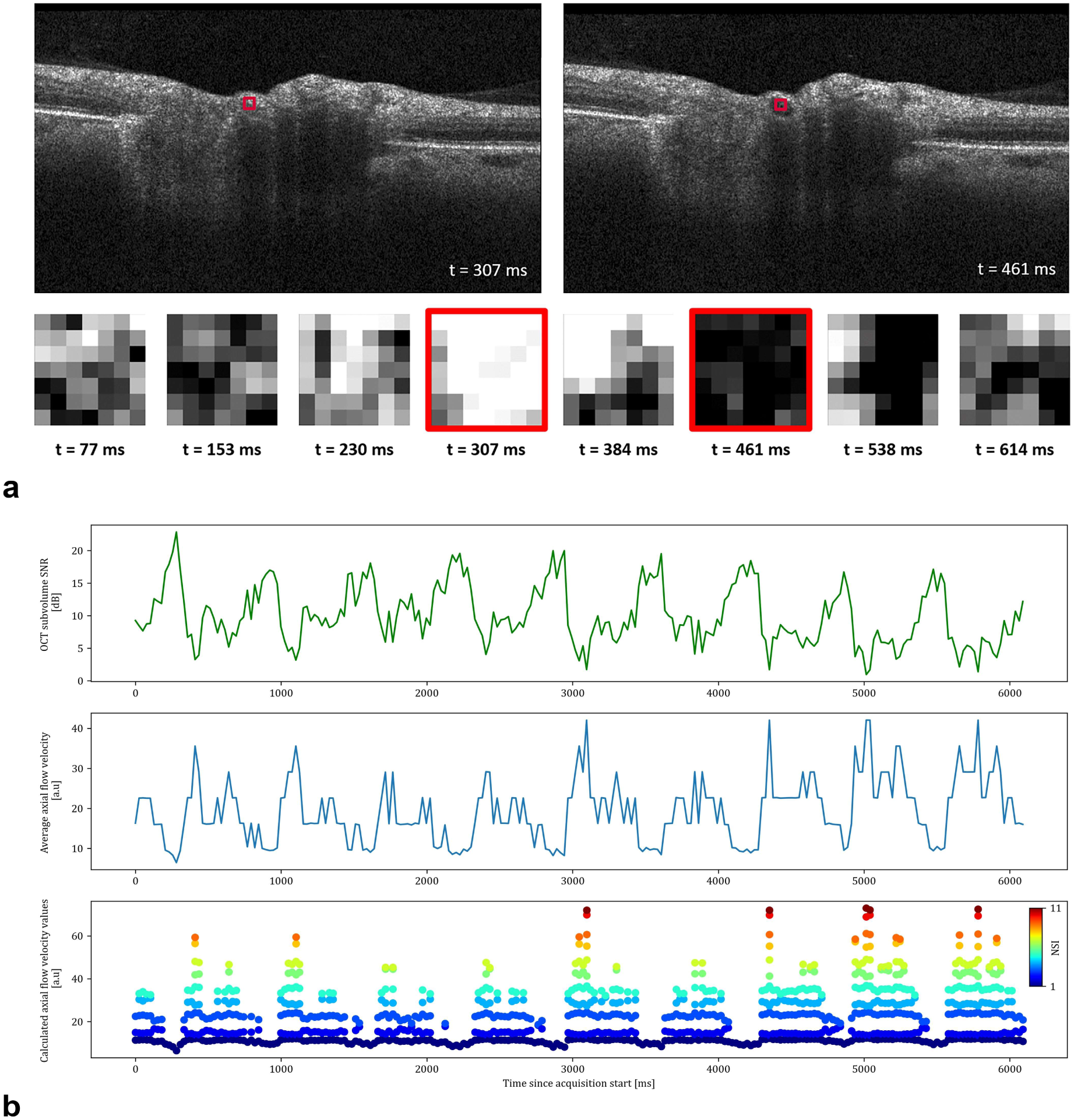
Time-resolved optical coherence tomography (OCT) B-scans of optic nerve head vessels. **a** Imaging was performed at the same location. Two B-scans and eight extracted 7 × 7 pixel arterial subvolumes (20 × 27 µm) are shown with timestamps. The corresponding subvolumes are framed in red, where the red frame on the original B-scan is enlarged for better visualisation. The intensity changes in the subvolume occur due to varying fringe washout of the arterial signal over time. **b** Analysis of retinal blood flow dynamics over 6 s in an artery with time-resolved OCT (A-scan integration time of 22.4 μs, inter-B-scan interval of ∼25 ms). Timestamp-matched mean SNR of the OCT subvolume (top row), average axial flow velocity (middle row) and calculated axial flow velocity values (bottom row) are presented. The average axial flow velocity line (middle row) represents the average of the numerical solutions, and the dots (bottom row) represent all numerical solutions to the SNR_Drop_ equation. dB: decibel, a.u.: arbitrary unit, NSI: numerical solution index.

**Figure 5.**
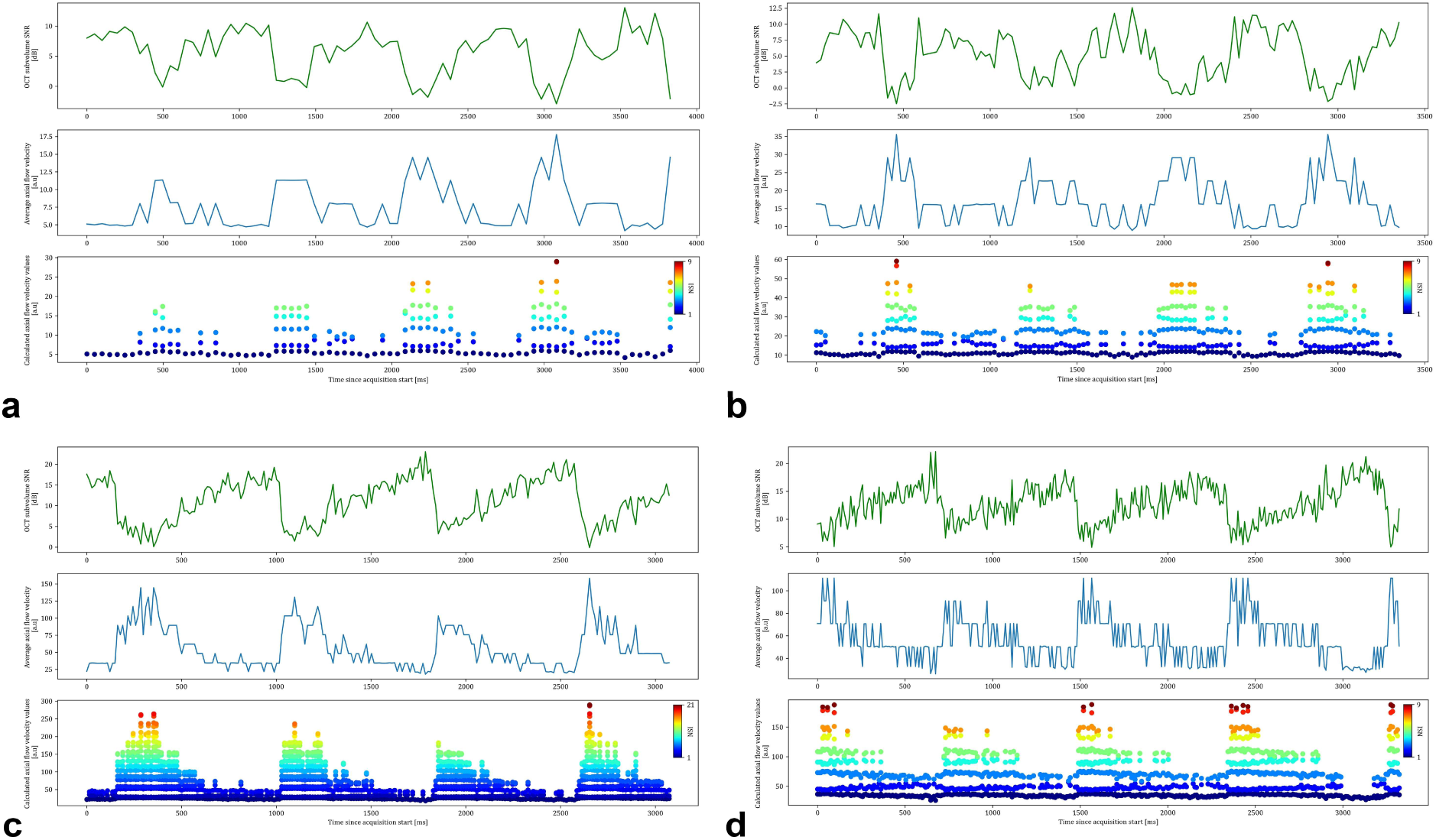
Calculated flow velocity profiles of time-resolved OCT intensity at the four different nominal acquisition rates (**a** 20 kHz, **b** 40 kHz, **c** 85 kHz and **d** 125 kHz). Each subfigure shows the acquisition duration in ms from the start on the x-axis. On the y-axes, the mean OCT SNR from the subvolume can be found on the top row, the average flow velocity (average of the numerical solutions to the SNR_Drop_ equation) in blue on the middle row and the calculated flow profiles (all numerical solutions) as dots on the bottom row.

When analysing the blood flow profiles at different eccentricities, pulsatility was evident close to the optic nerve centre. The pulsatility decreased towards the periphery, where a distinct change tended to occur when the vessel crossed the optic nerve head rim. Figure 6 provides a visual representation of this finding by showing the calculated blood flow profiles along with an en-face representation of their acquisition location. Further, we observed a decrease of the estimated average flow velocity towards the periphery, here again with an apparent decrease at the crossing of the vessel of the optic nerve head rim.

**Figure 6.**
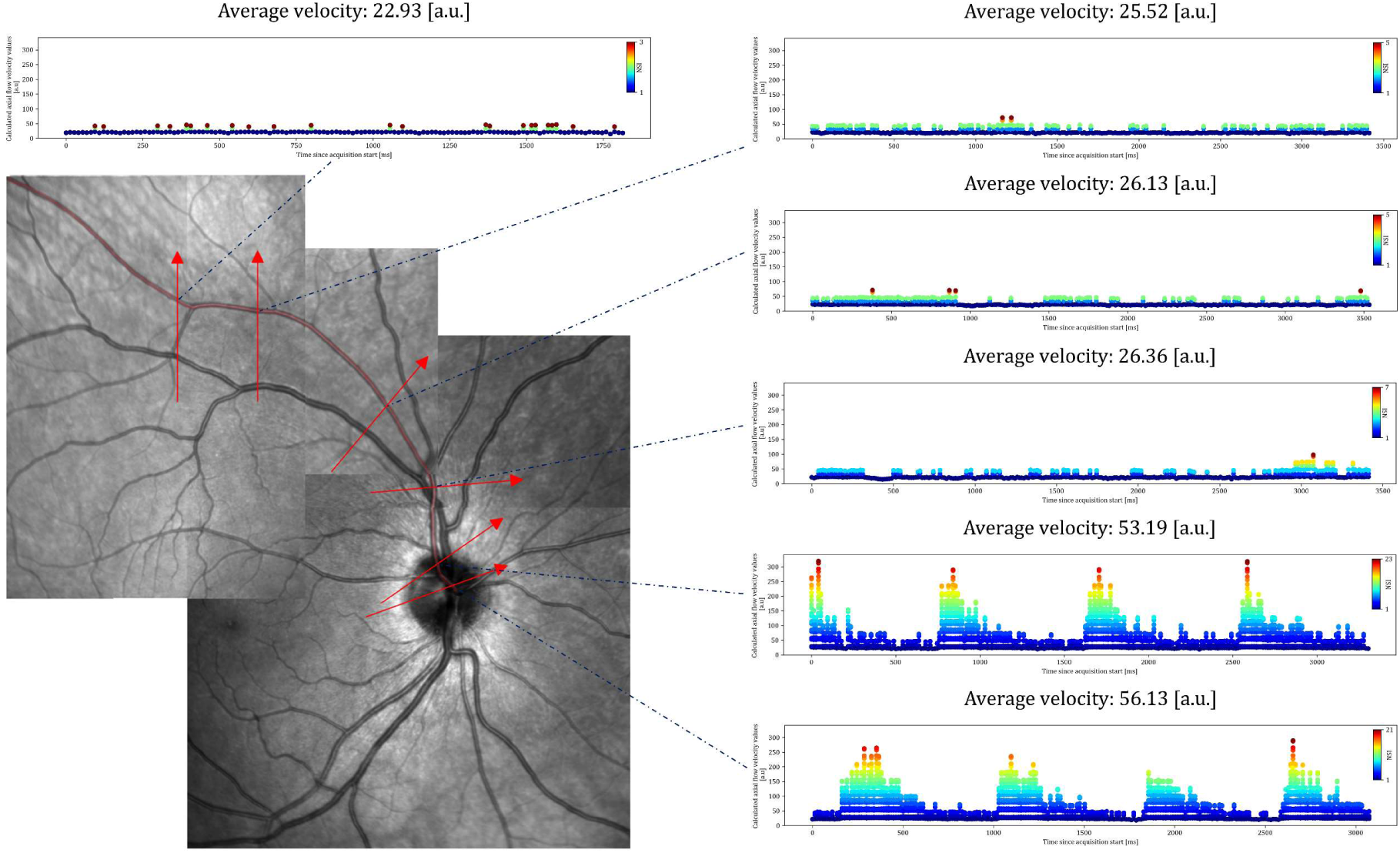
En-face representation of time-resolved acquisitions along an artery. The montage scanning laser ophthalmoscopy (SLO) of different acquisitions in the bottom left indicates the OCT acquisition locations as arrows, and the analysed artery is presented in transparent red. The corresponding time-resolved dynamic blood flow profiles show a clear pulsatility close to the optic nerve head centre, which diminishes beyond the optic nerve head rim. The estimated average blood flow velocities decrease from the optic nerve head centre towards the periphery.

### Quantitative analysis

We validated whether the SNR variability inside the vessels was different from the SNR variability in adjacent tissue, as the fringe washout variability could occur due to axial bulk motion. For this analysis, we annotated further retinal nerve fibre layer (RNFL) subvolumes in the proximity of an artery as the RNFL is known to have a high reflectivity in OCT.^27^ We analysed the SNR in the vessel (SNR_Vessel_), the SNR in the RNFL (SNR_RNFL_) and the SNR in the complete B-scan (SNR_B-scan_). The analysis confirmed that the SNR_Vessel_ was lower than the SNR_RNFL_. The SNR_B-scan_ was the highest as it was defined by the peak SNR in the complete image. The SNR_Vessel_ / SNR_B-scan_ ratio vessel clearly decreased with longer integration times while the SNR_RNFL_ / SNR_B-scan_ ratio showed less variability. This showed that the fringe washout inside the vessels was stronger at longer integration times. The SNR comparison of the artery and the RNFL are further described for the different integration times in Table 1. A visualisation of the fringe washout at the different integration times can be found in the Supplementary Video 3, where the SNR drops diminish with faster acquisitions.

**Table 1.** Vessel–RNFL comparison of SNR.

| Variable | 20 kHz, N = 78 | 40 kHz, N = 132 | 85 kHz, N = 228 | 125 kHz, N = 351 |
| --- | --- | --- | --- | --- |
| **SNR<sub>B-scan</sub> [dB]** | 39.7 (38.9, 40.6) | 37.8 (37.3, 38.5) | 36.0 (35.0, 36.9) | 32.6 (31.7, 33.8) |
| **SNR<sub>Subvolume</sub> [dB]** |  |  |  |  |
| Artery | 6.8 (2.6, 8.7) | 5.3 (2.1, 8.5) | 11.8 (6.2, 15.4) | 12.7 (10.0, 15.2) |
| RNFL | 33.7 (32.4, 34.6) | 32.2 (31.1, 33.4) | 30.7 (29.7, 31.6) | 28.7 (27.4, 29.6) |
| **Noise Level [RAW value]** | 55,186 (51,028, 61,463) | 110,620 (94,717, 124,661) | 202,044 (179,693, 228,765) | 146,278 (129,426, 165,785) |
| **SNR<sub>Subvolume</sub> / Peak SNR<sub>Subvolume</sub> [%]** |  |  |  |  |
| Artery | 4.68 (1.89, 7.09) | 2.76 (1.32, 5.79) | 1.40 (0.39, 3.27) | 2.23 (1.19, 3.96) |
| RNFL | 14.2 (10.7, 17.6) | 17.3 (13.3, 22.3) | 13.8 (10.9, 17.0) | 13.9 (10.5, 17.0) |
| **SNR<sub>Subvolume</sub> / SNR<sub>B-scan</sub> [%]** |  |  |  |  |
| Artery | 0.05 (0.02, 0.07) | 0.06 (0.03, 0.11) | 0.38 (0.13, 0.73) | 1.10 (0.58, 1.64) |
| RNFL | 24 (16, 30) | 27 (22, 34) | 28 (24, 33) | 42 (31, 48) |

Results are presented as median (IQR)
RNFL: retinal nerve fibre layer, N: number of B-scans with matched vessel–RNFL annotations, SNR: signal-to-noise ratio, SNR<sub>B-scan</sub>: SNR in the complete B-scan, SNR<sub>Subvolume</sub>: SNR in the B-scan subvolume, RAW value: value extracted from the raw .vol OCT files, Peak SNR<sub>Subvolume</sub>: maximal SNR on a pixel level in the complete subvolume over time, IQR: interquartile range.

An overview of the statistics and the calculated velocities per A-scan rate can be found in Tables 2 and 3. When analysing the absolute SNRs, SNR_B-scan_ and SNR_Vessel_ correlated with integration time. The calculated factors of the integration times of 44.8 μs at a nominal A-scan rate of 20 kHz, 22.4 μs at 40 kHz 11.2 μs at 85 kHz, 7.24 μs at 125 kHz are 6.19, 3.09, and 1.55 to 1 when taking the shortest integration time as a reference. Median SNR_B-scan_ factors are 5.83, 3.73, and 1.75 to 1, respectively. In summary, a longer integration time led to an increase of both absolute SNR_Vessel_ and SNR_B-scan_, and a decrease of the SNR_Vessel_ / SNR_B-scan_ ratio. This is also shown in the SNR_Subvolume_ / SNR_B-scan_ for the arteries in Table 1.

**Table 2.** Acquisition overview per nominal A-scan rate.

|  | 20 kHz, N = 109 | 40 kHz, N = 113 | 85 kHz, N = 122 | 125 kHz, N = 122 |
| --- | --- | --- | --- | --- |
| **B-scan Interval [ms]** | 49 (49, 49) | 25 (25, 25) | 13 (13, 13) | 9 (9, 9) |
| **Exam Duration [ms]** | 4,224 (3,528, 5,317) | 4,042 (3,582, 4,376) | 3,411 (2,950, 3,759) | 2,867 (2,059, 3,321) |
| **Annotated B-scans per Image Stack [n]** | 86 (72, 108) | 159 (141, 172) | 253 (219, 279) | 303 (218, 351) |
| **SNR<sub>B-scan</sub> [dB]** | 39.8 (37.3, 42.4) | 38.0 (35.3, 39.6) | 34.6 (31.0, 36.0) | 32.2 (29.9, 34.4) |
| **Peak B-scan Intensity [RAW value]** | 555,997,151 (340,536,329, 1,034,710,323) | 786,543,813 (345,832,675, 1,310,379,806) | 685,001,499 (347,938,689, 963,279,206) | 239,018,674 (168,368,268, 488,855,979) |
| **Annotated Image Stacks [N]** |  |  |  |  |
| Optic nerve head centre | 9 (8.3%) | 13 (12%) | 13 (11%) | 13 (11%) |
| NRM | 38 (35%) | 31 (27%) | 42 (34%) | 42 (34%) |
| 1 ODD from NRM | 16 (15%) | 17 (15%) | 17 (14%) | 17 (14%) |
| 2 ODDs from NRM | 14 (13%) | 19 (17%) | 17 (14%) | 17 (14%) |
| 3 ODDs from NRM | 17 (16%) | 17 (15%) | 17 (14%) | 17 (14%) |
| 4 ODDs from NRM | 12 (11%) | 13 (12%) | 13 (11%) | 13 (11%) |
| 5 ODDs from NRM | 3 (2.8%) | 3 (2.7%) | 3 (2.5%) | 3 (2.5%) |

Results are presented as median (IQR) of the average values per image stack | N (% of image stacks per acquisition rate)
N: number of time-resolved image stacks, SNR<sub>B-scan</sub>: signal-to-noise ratio in the complete B-scan; RAW value: value extracted from the raw .vol OCT files; NRM: Neuroretinal rim, 1 ODD from NRM: eccentricity of 1 optic disc diameter from the neuroretinal rim, IQR: interquartile range.

**Table 3.**
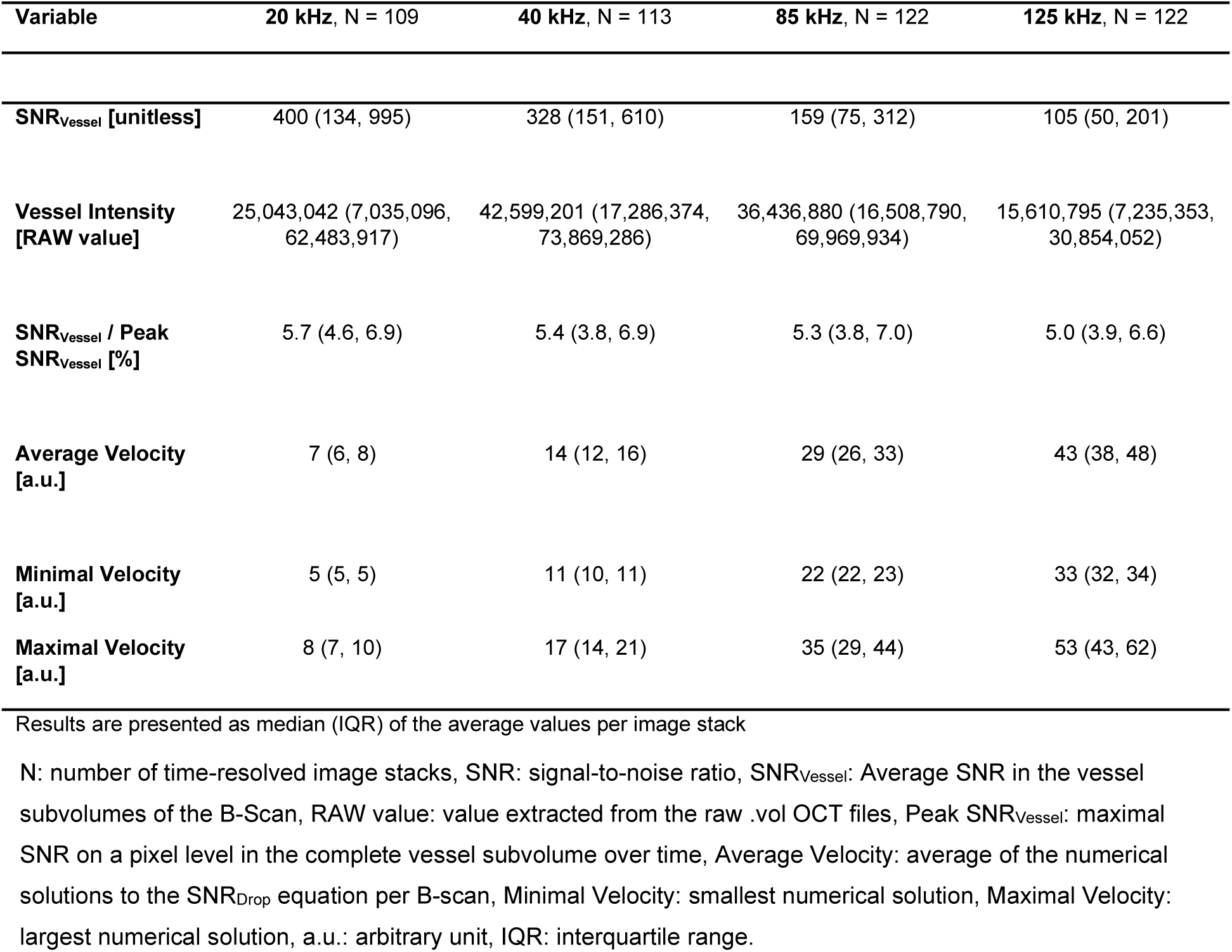
Vessel subvolume SNRs and calculated velocities.

| Variable | 20 kHz, N = 109 | 40 kHz, N = 113 | 85 kHz, N = 122 | 125 kHz, N = 122 |
| --- | --- | --- | --- | --- |
| **SNR<sub>Vessel</sub> [unitless]** | 400 (134, 995) | 328 (151, 610) | 159 (75, 312) | 105 (50, 201) |
| **Vessel Intensity [RAW value]** | 25,043,042 (7,035,096, 62,483,917) | 42,599,201 (17,286,374, 73,869,286) | 36,436,880 (16,508,790, 69,969,934) | 15,610,795 (7,235,353, 30,854,052) |
| **SNR<sub>Vessel</sub> / Peak SNR<sub>Vessel</sub> [%]** | 5.7 (4.6, 6.9) | 5.4 (3.8, 6.9) | 5.3 (3.8, 7.0) | 5.0 (3.9, 6.6) |
| **Average Velocity [a.u.]** | 7 (6, 8) | 14 (12, 16) | 29 (26, 33) | 43 (38, 48) |
| **Minimal Velocity [a.u.]** | 5 (5, 5) | 11 (10, 11) | 22 (22, 23) | 33 (32, 34) |
| **Maximal Velocity [a.u.]** | 8 (7, 10) | 17 (14, 21) | 35 (29, 44) | 53 (43, 62) |

Results are presented as median (IQR) of the average values per image stack
N: number of time-resolved image stacks, SNR: signal-to-noise ratio, SNR<sub>Vessel</sub>: Average SNR in the vessel subvolumes of the B-Scan, RAW value: value extracted from the raw .vol OCT files, Peak SNR<sub>Vessel</sub>: maximal SNR on a pixel level in the complete vessel subvolume over time, Average Velocity: average of the numerical solutions to the SNR<sub>Drop</sub> equation per B-scan, Minimal Velocity: smallest numerical solution, Maximal Velocity: largest numerical solution, a.u.: arbitrary unit, IQR: interquartile range.

The SNR_Vessel_ / Peak SNR_Vessel_ ratio corresponds to the SNR drop with which the flow velocities were calculated. This ratio remained constant across the varying integration times. Given that Yun et al.’s model for flow velocities is dependent on the integration time, the calculated flow velocities showed a linear relationship with their integration time ratio.^18^ In the analysis of the flow velocities at different distances from the optic nerve head of the complete cohort, decreasing calculated mean flow velocity along the vessel arch were observed. A summary overview of the flow velocities along the vessel arch is shown in Figure 7, with corresponding examples of SLOs.

**Figure 7.**
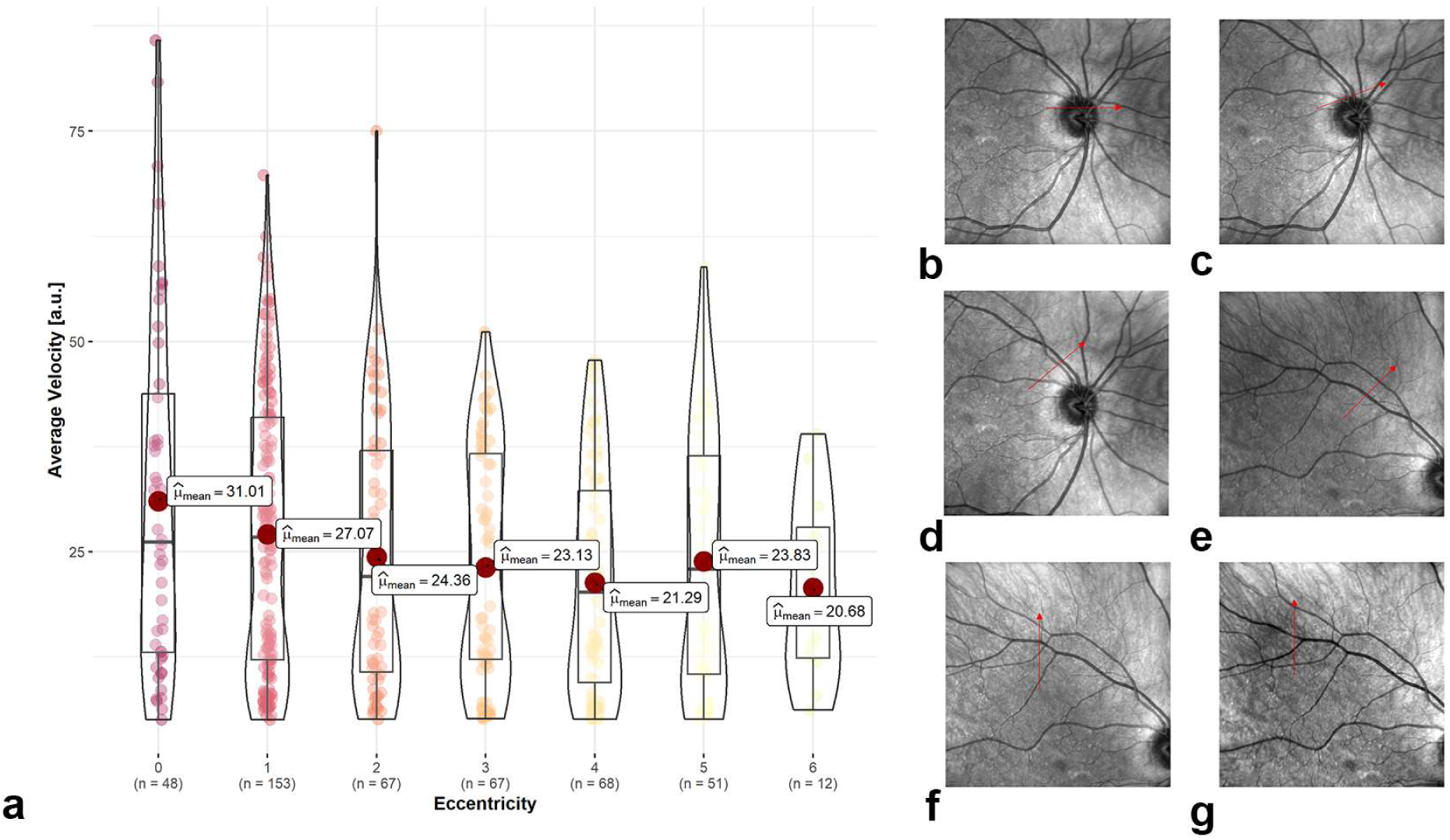
**a** Estimated average blood flow velocity of the measured vessels at different eccentricities represented as violin-boxplots and with the means highlighted as red dots. **b–g** Example scanning laser ophthalmoscopy (SLO) acquisitions with the positioning of the continuous B-scans highlighted by a red arrow corresponding to eccentricities 0–5. **b** ≙ 0: Optic nerve head centre, **c** ≙ 1: Neuroretinal rim, **d** ≙ 2: eccentricity of 1 optic disc diameter (ODD) from the neuroretinal rim, **e** ≙ 3: eccentricity of 2 ODDs from the neuroretinal rim, **f** ≙ 4: eccentricity of 3 ODDs from the neuroretinal rim, **g** ≙ 5: eccentricity of 4 ODDs from the neuroretinal rim.

## Discussion

To analyse the dynamics within retinal structures and, more specifically, in vessels, we developed acquisition protocols for dynamic time-resolved OCT measurements utilising a commercially available OCT device widely used in clinics. Taking advantage of the fringe washout phenomenon of spectral-domain OCTs, we calculated time-resolved velocity profiles based on the SNR drop within the vessels. In addition, we found pulsatile blood flow profiles, where the amplitude and average velocity decreased from the centre of the optic nerve head towards the retinal periphery.

This study presents several novel possibilities for the analysis of OCT data acquired with a commercially available device. First, time-resolved dynamic OCT with a matching of B-scans to a time axis allowed us to assess structural changes with an interval of as low as 9 ms between B-scans. The matching of the B-scans to a time axis allowed for generating videos with real interframe intervals (Supplementary Videos 1 and 2). Considering that inter-B-scan intervals differ when tracking is activated and individual B-scans are averaged, continuous visualisations of B-scans can create an incorrect impression of the speed at which changes happen.

Second, this study presents methods for comparing OCT acquisitions with different integration times. As OCT phenomena, such as the fringe washout, are affected by the integration time, it is important to consider parameters such as the image quality or the OCT intensity, which vary with different integration times.^18,28^ In this study, we showed in vivo that the fringe washout phenomenon varies for different integration times. The fringe washout in vessels was stronger at longer integration times, which should be considered in further studies.

Third, we combined this knowledge to investigate and visualise pulsatile dynamics within vessels in structural OCT. For this, we took advantage of the fact that the fringe washout analysis allows for the calculation of corresponding flow velocities. We generated blood flow profiles by calculating the flow velocity of the axial motion component for each time-resolved B-scan. OCT is primarily utilised for visualising retinal tissue, but it provides several advantages over OCTA in certain scenarios. In particular, structural OCT allows to directly work with the raw intensities from the acquisition as opposed to post-processed images. As a result, OCT can also be advantageous for the analysis of vessels, as it provides direct information about the characteristics of the tissue being imaged.

The analysis of the fringe washout would generally allow the calculation of velocities in mm/s. However, there are several limitations to quantitative values that we could not address in this work. The fringe washout is composed of axial and lateral components. Weighing both components is possible if the angle of the vessel and sample motion is known.^29^ However, the axial component is larger by orders of magnitude compared with the lateral component in the case of oblique motion.^19^ A further limitation that must be addressed is the reference intensity to calculate the SNR drop, which cannot be determined in a moving system. The optical properties of the eye, such as the clarity of the optical media as the lens, the cornea or even the momentaneous tear film affect the SNR.^30^ Furthermore, the peak SNR inside a vessel is also affected by fringe washout, as there is permanent motion in a larger vessel in vivo. Hence, an absolute reference value or an external calibration seems difficult to determine, even under ideal circumstances. Further, the sensitivity roll-off along the A-scan axis was not assessed and hence not included in the noise calculations. An analysis of the transformation matrices used in the registration process could assist in calculating the fringe washout component caused by bulk eye motion. In this work, we focused on the analysis of the axial component of the fringe washout to calculate potential blood flow velocities and estimate relative flow profiles in vivo. For this, we calculated numerical solutions to the SNR_Drop_ equation representing potential axial flow velocities. We present estimated flow velocity profiles in arbitrary units as an average of all the numerical solutions per B-Scan. It’s important to note that these velocity profiles are estimates as there are multiple numerical solutions to the SNR_Drop_ equation. Potentially, for certain B-scans a lower numerical solution could represent the real velocity, while for others with the same SNR_Drop_ a higher numerical solution would be correct. To determine which numerical solutions to the equation accurately reflect the true axial flow velocity values, further validation of the technique is required.

An objective reference value could help align the different flow velocities we found at different acquisition rates by using the formula by Yun et al.^18^ The calculated average, minimum and maximum flow velocities seem to be inversely dependent on the integration time as shown in Table 3. Further, we found that the absolute SNR in the vessels increases with increasing integration time, however, that the relative SNR drop within the vessels remained constant across the integration times. The major advantage of this finding is that blood flow profiles can be estimated and visualised independently from the integration time, even at higher acquisition rates where there is less relative fringe washout.

The decreasing velocity of the flow profiles along the vessel arch suggest that the method can grasp decreasing velocities in accordance with known physiologic principles. However, this cannot yet be supported by our data as the angle between the beam and the flow direction varies across the retina. Such changes in the angle also affect the axial velocity component. In accordance with this, we noticed that the pulsatility of the calculated blood flow profiles tends to decrease when an artery crosses the optic nerve head rim. For a precise measurement of the flow velocity, again the lateral component or the angle of the vessel would have to be known.^29^ Other techniques to measure retinal blood flow exist, such as Doppler OCT, laser speckle flowgraphy and VISTA in OCTA.^5,6,8,31^ The analysis of retinal blood flow based on intensity changes has also been presented for a neural network-based approach.^32^ However, these technologies are mainly used in research environments and are not yet regularly in clinical practice as the required phase information or A-scan acquisition rates are not widely available. Our method allows to calculate retinal blood flow velocity profiles with a widely used clinical device while at the same time contributing to bridge the gap left by other measurement techniques. Techniques such as laser Doppler flowmetry can only be used if the Doppler angle is known; laser speckle flowgraphy does not allow for a depth-resolved flow measurement; and VISTA is an extension of OCTA that has been presented for the macular area only.^8,31^ This could be mainly due to the fact that the line rates of the lasers are currently not high enough to measure changes in the saturation of signal changes for VISTA outside the macular area. With further validation, our method could allow the visualisation of depth- and time-resolved flow profiles, as shown in this study for vessels around the optic nerve head. Ideally, our method could be further enhanced by including Doppler angle and phase shift information to translate combined findings into clinical practice.^5,10,19^ A validation of newly obtained blood flow profile values could be achieved by comparing them with measurements from flow phantoms.^19^

To analyse retinal blood flow is relevant as the vasculature is involved in many ocular diseases, such as diabetic retinopathy and glaucoma. Blood flow disturbances, such as haemorrhage or ischemic attacks, can lead to vision loss and identifying vessels at risk could help prevent vascular injuries. For accurate and easy-to-use measurements, blood flow measurements must be implemented in clinics. We aim to contribute to this goal with the proposed time-resolved structural OCT-generated flow profiles. Thus far, our method has only been tested in healthy participants and not yet in subjects with ocular diseases. The short-term reproducibility for the generation of flow profiles at the same location was good. Very similar flow profiles could be generated during acquisitions with different integration times at the same location. Short-term reproducibility at the same location with the same integration time as well as inter-visit reproducibility and variability still need to be determined. The method could be further validated by imaging subjects with diseases such as glaucoma or diabetic retinopathy, as their blood flow has been shown to be altered with other measuring techniques.^33–35^ The new estimates of retinal blood flow profiles could also be compared against the previously described techniques in further studies.

Future work will include a deeper analysis of the pulsatile flow profiles. As shown in Figures 4 and 5 and Supplementary Videos 2 and 3, the pulse wave appeared to have a bimodal peak, which would be congruent with the propagation of the aortic pulse wave.^36^ In addition, the pulse wave analysis, in combination with absolute velocity and flow information, could also be used to analyse the retinal blood flow volume and parts of the cardiac output arriving at the eye from the heart.^10,37^ An extension of this could also include the investigation of the best acquisition location, differences across branching points and general differences between arteries and veins. The current measurements are consecutive B-scan acquisitions over time, with a minimal inter-B-scan interval of ∼9 ms at 1024 A-scans. With fast acquisition speeds and a small number of B-scans, 4D cross-sectional acquisitions can become feasible. Hereby, the pulsatile nature of the fringe washout indicates a correlation with the cardiac cycle and a representation of the pulse propagation in the vasculature of the optic nerve head. In vivo measurements could investigate the validity of models about the pulse propagation towards the eye.^38^

## Conclusions

In summary, this study showed that time-resolved OCT acquisitions are feasible in vivo and that continuous acquisitions over several seconds can be made with a widely used clinical device. We showed that dynamic blood flow profiles can be calculated from time-resolved OCT acquisitions. Time-resolved dynamic OCT, with its high spatial and temporal resolution, holds promising information that can be further investigated as a novel clinically applicable parameter for the assessment of retinal blood flow velocity profiles.

## Data Availability

The data presented in this paper are not publicly available due to data protection regulations. Interested parties may request access to the data from the corresponding author (PV) upon reasonable request and approval from the concerned institutional review boards.

## Acknowledgements

The authors wish to thank the Swiss National Science Foundation, the Janggen-Pöhn-Stiftung and the AlumniMedizin Basel for their financial support of this project. In addition, the researchers would like to thank all the participants who volunteered for this study.

## Supplementary Material

**Supplementary Video 1**

Time-resolved dynamic optical coherence tomography of an artery (cross-sectional) at the centre of the optic nerve head.

**Supplementary Video 2**

Time-resolved dynamic optical coherence tomography of an artery (longitudinal) at the centre of the optic nerve head.

**Supplementary Video 3**

Comparison of time-resolved dynamic optical coherence tomography with corresponding blood flow velocity profiles at four different integration times.

